# Expanding the genetic landscape of endometriosis: Integrative -omics analyses implicate key genes and pathways in a multi-ancestry study of over one million women

**DOI:** 10.1101/2024.11.26.24316723

**Authors:** Lindsay A Guare, Jagyashila Das, Lannawill Caruth, Ananya Rajagopalan, Alexis T. Akerele, Ben M Brumpton, Tzu-Ting Chen, Leah Kottyan, Yen-Feng Lin, Elisa Moreno, Ashley J Mulford, Marija Simona Dombrovska, Yuan Luo, Vita Rovite, Alan R Sanders, Craig Teerlink, Danielle Candelieri, Noemie Elhadad, Andrew Hill, Gail P. Jarvik, James Jaworski, Julie Lynch, Shinichi Namba, Yukinori Okada, Yue Shi, Yuya Shirai, Jonathan Shortt, Wei-Qi Wei, Chunhua Weng, Yuji Yamamoto, Penn Medicine Biobank, Regeneron Genetics Center, Global Biobank Meta-analysis Initiative, Sinead Chapman, Wei Zhou, Kyong-Mi Chang, Todd Edwards, Suneeta Senapati, Digna R. Velez Edwards, Shefali Setia-Verma

## Abstract

We report the findings of a genome-wide association study (GWAS) meta-analysis of endometriosis across 14 biobanks worldwide, including 32% non-European patient participants, as part of the Global Biobank Meta-Analysis Initiative (GBMI). Eleven meta-analyses detected 58 total associated regions (27 previously unreported), including six yielded by imaging- and surgery-confirmed phenotypes. We detected the first genome-wide significant loci uniquely driven by the African-ancestry (1q42.*13, 2p13*.*3*, and *20q13*.*2*) and Admixed-American-ancestry (*21q22*.*13*) meta-analyses. Leveraging our large and diverse study population, we observed SNP heritability estimates of 9-13% for all ancestry groups, and 14 regions had at least one variant in the credible set after fine-mapping. Investigating the complex array of endometriosis comorbidities and risk factors revealed 135 genetically correlated phenotypes and 95 with evidence of vertical pleiotropy, including triglycerides and anxiety disorders. We prioritized 35 disease-relevant cellular contexts from the endometrial cell atlas and found 322 examples of differentially expressed genes in cells from donors with endometriosis. Including further high-throughput multi-omic analyses, we have implicated a total of 314 genes in endometriosis pathogenesis. This diverse, comprehensive GWASs, with downstream analyses spanning molecular to phenotypic scales, provide detailed evidence for aspects of endometriosis including the role of immune cell types and cellular proliferation. These interconnected pathways and risk factors underscore the complex, multi-faceted etiology of endometriosis, suggesting multiple targets for precise and effective therapeutic interventions.

## Introduction

Endometriosis, a debilitating, chronic, and systemic condition, characterized by the growth of endometrial-like tissues outside of the uterus, affects approximately 10% of women of reproductive age worldwide^1,2^. The precise mechanisms of these lesions are presently unknown. Endometriosis is incurable, leaving patients to manage severe pain^3^ and cope with fertility challenges^4^. In addition to its reproductive burden, growing epidemiological and genetic evidence indicates a link between endometriosis and cardiovascular disease risk, suggesting that shared inflammatory and vascular mechanisms may contribute to both conditions^5–9^. Despite its prevalence and impact, the etiology of endometriosis remains poorly understood, hampering efforts to develop effective diagnostic tools and targeted treatments.

Genome-wide association studies (GWASs) have been an important tool for suggesting candidate genes and pathways linked with endometriosis^10–12^. GWASs for endometriosis have uncovered over 40 loci associated with endometriosis risk. However, like most GWASs historically, the previous endometriosis studies have primarily included European ancestry populations and at a smaller scale, East-Asian^13^. While the broad sense heritability of endometriosis has been estimated as 47% via a twin study^14^, previous endometriosis GWASs have only explained about 7% of phenotypic variance with common variants^10,11^ and have fallen short in elucidating the full spectrum of its pathophysiology.

The advent of large-scale biobanks has revolutionized genetic research, offering unique opportunities to conduct well-powered studies across diverse populations^15–18^. As more diverse genomic datasets become available like the Penn Medicine Biobank (PMBB)^19^, the All of Us Research Program (AOU)^20^, and the Million Veterans Program (MVP)^21^, they enhance the discovery of trait loci through increased statistical power and variation enriched in non-European populations^15^. A worldwide consortium of genomic researchers, the Global Biobank Meta-Analysis Initiative (GBMI), has been established to facilitate collaboration in GWAS studies on unprecedented scales. In this study, we at the PMBB have collaborated with 12 other biobanks across several countries^20,22–32^ and incorporated publicly-available summary statistics from FinnGen, totaling 14 global datasets. These resources not only enhance statistical power to detect novel associations but also provide rich phenotypic data, enabling more refined analyses and the investigation of pleiotropy across related traits and conditions. Such large-scale, multi-phenotype datasets offer a unique opportunity to better understand shared genetic architecture between endometriosis and other complex diseases. Moreover, the inclusion of diverse ancestries in genetic studies is crucial to aid in fine-mapping and for improving the generalizability of findings and addressing health disparities. This is particularly pertinent for endometriosis, where significant variations in prevalence and clinical presentation have been observed across different ethnic groups^33^.

Beyond GWAS, there are many methods which can be employed *in silico* to investigate other elements of the central dogma, providing further validation of GWAS loci and discovery of additional disease-associated genes. Recent advances in multi-omics technologies have further expanded our ability to translate genetic associations into biological insights. Integration of GWAS results with multi-omic QTL and single-cell datasets using methods such as imputed association studies and Mendelian randomization allows for a more comprehensive understanding of disease mechanisms, potentially identifying novel therapeutic targets and biomarkers. For complex diseases like endometriosis, where the interplay between genetic predisposition and tissue-specific processes is central, integrative approaches across multiple layers of biology are valuable^34–36^ . Given these considerations, there is a critical need for a large-scale, ancestrally diverse GWAS of endometriosis that incorporates rigorous phenotyping and leverages multi-omics data to provide a more complete picture of the disease’s genetic landscape and underlying biology.

Here, we present the results of a large-scale GWAS meta-analysis that significantly advances our understanding of endometriosis pathophysiology and sheds light on shared biological mechanisms underlying comorbid conditions. Our study encompasses a diverse cohort from 14 biobanks worldwide, with over 30% non-European participants, enhancing the generalizability of our findings. We employed a comprehensive phenotyping approach, including not only a wide spectrum of endometriosis presentations but also surgically confirmed and procedure-validated (surgery or imaging) phenotypes. This robust phenotyping strategy enables greater resolution when examining the genetic architecture of endometriosis. Through high-throughput exploration of pleiotropy and integrative multi-omics analyses, we have explored numerous layers of endometriosis pathogenesis. These findings provide unprecedented insights into the risk factors and molecular mechanisms driving this complex disorder, paving the way for improved diagnostics and targeted therapeutic interventions.

## Results

### EHR-Based Phenotyping of Endometriosis in Genetically Diverse Datasets

Of the six case-control definitions (W - <u>W</u>ide, Wex - <u>W</u>ide <u>e</u>xcluding adenomyosis, PCN.v2 and PCN.v2 - <u>P</u>rocedure-<u>C</u>onfirmed <u>N</u>arrow [vs (1) all and (2) confirmed controls], and SCN.v1 and SCN.v2 - <u>S</u>urgically-<u>C</u>onfirmed <u>N</u>arrow [vs (1) all and (2) confirmed controls]), the largest phenotype, W, comprised 1,063,657 women (31.9% non-EUR), including 50,265 cases. The Wex phenotype, for which several biobanks were omitted, had a total sample size of 615,050 (N_cases_ = 12,262). Sample sizes for the PCN and SCN phenotypes were smaller (Table 1), with summary statistics included from only nine of the 14 biobanks (Supplementary Table 1).

**Table 1:** All GWAS meta-analysis sample sizes, with the 11 main ones performed shaded in gray. All six phenotypes had a multi-ancestry meta-analysis as well as single-ancestry meta-analyses for AFR, AMR, CSA+SAS, EAS, and EUR. There were not enough CSA+SAS or EAS studies to conduct meta-analyses for all the narrow phenotypes. Phenotype abbreviations: W = wide, Wex = wide excluding adenomyosis, PCN = procedure-confirmed narrow, SCN = surgically-confirmed narrow, .v1 = versus all controls, .v2 = versus confirmed controls. Wide phenotype definitions utilized endometriosis diagnosis codes while narrow definitions combined diagnoses with procedure history (i.e. laparoscopy, non-obstetric ultrasound). Ancestry abbreviations: AFR = African, AMR = admixed American, CSA = central/south Asian, SAS = south Asian, EAS = east Asian, EUR = European.

| Phenotype | Ancestry | N | Cases | Prevalence | N effective |
| --- | --- | --- | --- | --- | --- |
| W | AFR | 96,012 | 4,289 | 4.47% | 16,390 |
|  | AMR | 67,335 | 2,136 | 3.17% | 8,273 |
|  | CSA+SAS | 8,164 | 259 | 3.17% | 1,003 |
|  | EAS | 168,209 | 7,833 | 4.66% | 29,873 |
|  | EUR | 723,937 | 35,748 | 4.94% | 135,931 |
|  | META | 1,063,657 | 50,265 | 4.73% | 191,559 |
| Wex | AFR | 92,202 | 1,575 | 1.71% | 6,192 |
|  | AMR | 66,219 | 1,020 | 1.54% | 4,017 |
|  | CSA+SAS | 5,066 | 62 | 1.22% | 245 |
|  | EAS | 86,667 | 1,584 | 1.83% | 6,220 |
|  | EUR | 364,896 | 8,021 | 2.20% | 31,379 |
|  | META | 615,050 | 12,262 | 1.99% | 48,070 |
| PCN.v1 | AFR | 70,959 | 1,152 | 1.62% | 4,533 |
|  | AMR | 58,802 | 750 | 1.28% | 2,962 |
|  | EAS | 78,999 | 155 | 0.20% | 619 |
|  | EUR | 317,481 | 4,053 | 1.28% | 16,005 |
|  | META | 526,241 | 6,110 | 1.16% | 24,156 |
| PCN.v2 | AFR | 12,958 | 1,152 | 8.89% | 4,198 |
|  | AMR | 9,472 | 750 | 7.92% | 2,762 |
|  | EAS | 1,539 | 155 | 10.07% | 558 |
|  | EUR | 56,134 | 4,053 | 7.22% | 15,041 |
|  | META | 80,103 | 6,110 | 7.63% | 22,576 |
| SCN.v1 | AFR | 70,308 | 501 | 0.71% | 1,990 |
|  | AMR | 58,477 | 425 | 0.73% | 1,688 |
|  | EUR | 284,932 | 2,573 | 0.90% | 10,199 |
|  | META | 413,717 | 3,499 | 0.85% | 13,878 |
| SCN.v2 | AFR | 1,912 | 501 | 26.20% | 1,479 |
|  | AMR | 2,291 | 425 | 18.55% | 1,385 |
|  | EUR | 14,147 | 2,573 | 18.19% | 8,420 |
|  | META | 18,350 | 3,499 | 19.07% | 11,327 |

The case rates varied across a wide range for the different phenotype definitions, from as low as 0.2% (EAS PCN.v1) to as high as 26.20% (AFR SCN.v2). Z-tests for proportion comparisons showed that EUR has a significantly higher proportion of cases than all other ancestries for the W and Wex phenotypes. For PCN.v2 and SCN.v2, however, EUR demonstrated the lowest prevalence (significantly lower than EAS for PCN.v2 and AFR for SCN.v2). EAS had the lowest case rate for PCN.v1. The rate of procedure or surgical confirmation (PC or SC) among controls was relatively similar amongst ancestry groups except for EAS: AFR – 16.91% PC; 2.02% SC, AMR – 15.02% PC; 3.21% SC, EAS – 1.76% PC, EUR – 16.62% PC, 4.10% SC.

### Multi-Ancestry Wide Phenotype GWAS Meta-Analysis Results

Across the 11 meta-analyses (six multi-ancestry meta-analyses for the different phenotype definitions plus five single-ancestry W meta-analyses), we identified 3,342 unique genome-wide significant variants (P < 5 × 10^−8^), corresponding to 5,332 associations (Supplementary Table 2) which replicate associations of 65 out of 177 variants in the GWAS catalog (Supplementary Table 3). Relative to previous large GWASs, we achieved moderate to high replication: *Sapkota et al 2017* (20 / 21 significant), *Sakaue et al 2021* (6 / 9 significant), *Rahmioglu et al 2023* (25 / 46 significant; +6 nominal), *Jinn-Chyuan Sheu et al 2024* (11 / 16 significant), *Gualdo et al 2025* (12 / 13 significant).

These associations were aggregated into 249 linkage disequilibrium (LD) clumps (Supplementary Table 4), which further merged into 108 distinct loci across the 11 meta-analyses (Supplementary Table 5). In total, 143 independent signals were detected by conditioning analysis, with eight loci containing more than one signal, including 25 and 11 near the *SYNE1* locus for the EUR and META W GWASs, respectively (Supplementary Table 6). When we consolidated loci shared between meta-analyses, we arrived at 58 total regions. Region highlighted in Figure 1 are labeled with the most significant corresponding gene from the downstream multi-omic analyses (described below) or the cytoband (Supplementary Table 7). There were 44 multi-ancestry endometriosis-associated regions. Besides those, there were three EUR-specific W regions (*GRB14, ZNF536*, and *CHD6*) and two AFR-specific W region (*2p13.3* and *20q13.2*), totaling 49 W regions annotated in Figure 1a.

**Figure 1:**
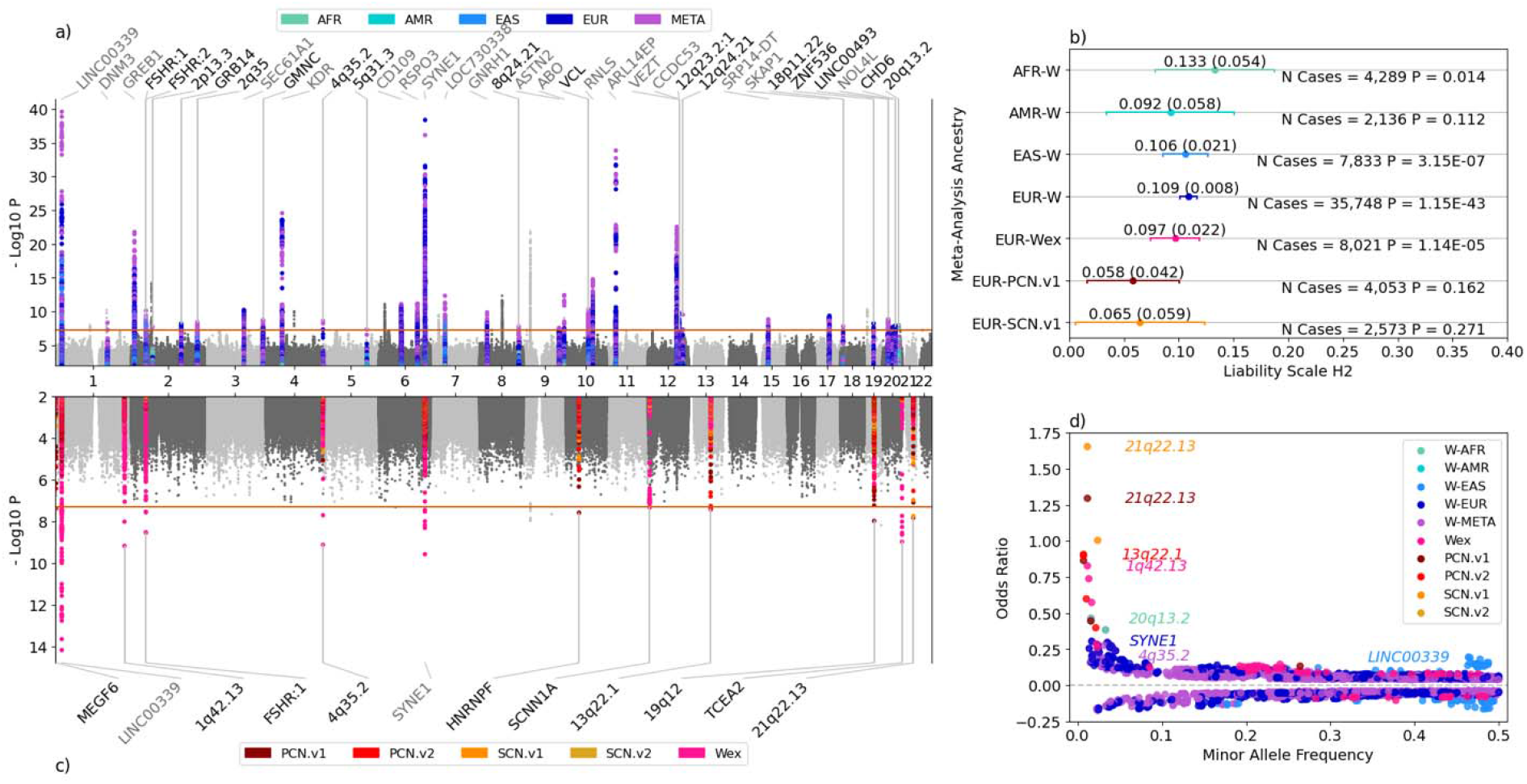
Overview of 11 GWAS meta-analysis results. a) Top left: overlayed Manhattan plots for the five <u>W</u>ide endometriosis phenotype GWASs (multi-ancestry + four single-ancestries). All significant regions are annotated with black text highlighting the previously unreported hits. b) Top right: LD-score regression (using LDSC) heritability estimates for the W GWASs; AMR is excluded because there was not sufficient sample size to use LDSC. c) Bottom left: Overlayed Manhattan plots for the five multi-ancestry GWASs for the other phenotypes. d) Bottom right: odds ratio versus minor allele frequency with the largest effect for each GWAS highlighted. Phenotype abbreviations: W = wide, Wex = wide excluding adenomyosis, PCN = procedure-confirmed narrow, SCN = surgically-confirmed narrow, .v1 = versus all controls, .v2 = versus confirmed controls. Ancestry abbreviations: AFR = African, AMR = admixed American, EAS = east Asian, EUR = European.

Based on the test with the strongest p-value for the lead SNPs in those 49 regions, 30 showed low heterogeneity, 17 moderate, one large (*2p*13.3, I^2^ = 60%), and one very large (*SYNE1*, I^2^ = 79%) (Supplementary Table 7). When comparing the lead SNPs in the regions with moderate or greater heterogeneity, four of them demonstrate non-overlapping effect size 95% intervals between two ancestries: *DNM3* (EUR 1.02-1.06 vs SAS 1.08-1.55), *2q35* (EUR 1.02-1.06 vs EAS 1.07-1.16), *LOC730338* (EAS 0.81-0.91 vs EUR 0.94-0.97), and *CCDC53* (EUR 0.93-0.96 vs AFR 1.01-1.10). Of the 49 unique W regions, 31 of them contained SNPs reported to have associations with endometriosis in the GWAS catalog^37^. The associations between endometriosis and the 18 remaining W regions are reported here for the first time.

Liability scale genome-wide SNP heritability estimates and standard errors (h^2^) from the W single-ancestry meta-analyses are shown in Figure 1b. The h^2^ estimates and their standard errors were: 0.133 (0.054) for AFR, 0.106 (0.021) for EAS, and 0.108 (0.008) for EUR. Heritability estimates (via the LD-score regression tool, LDSC) for all analyses are provided in Supplementary Table 8. Partitioned LD-score regression (Supplementary Table 9) attributed significant proportions of the phenotypic variance to enhancers active in uterine tissue (epigenome map state called “EnhA2”) in EAS (prop_h2_ = 33%, enrichment = 26-fold, P_prop h2_ = 1.2 × 10^−3^) and EUR (prop = 20%, enrichment = 15-fold, P = 6.6 × 10^−4^). Two others explained significant portions of the heritability: ncRNA_gene (non-coding RNA genes) in EAS (prop_h2_ = 21%, enrichment = 1.4-fold, P = 2.6 × 10^−3^) and TssFlnk (regions flanking transcription start sites) in EUR (prop_h2_ = 7%, enrichment = 1.3-fold, P_prop h2_ = 0.031). Cross-ancestry genetic correlation estimates with Popcorn were non-significant, with none being significantly different from one, likely due to sample size limiting the strength of individual heritability estimates^38,39^. The strongest estimate was between EAS and EUR: ρ_gi_ = 0.76, P = 0.064 (all genetic impact correlation estimates are available in Supplementary Table 10).

### Precise Phenotype Analyses Replicate Known Loci and Reveal Additional Loci

Across the remaining phenotype meta-analyses, we identified 14 significant regions: eight with Wex, three with just PCN.v1, one with both PCN.v1 and SCN.v1, and two with PCN.v2. Ten lead SNPs demonstrated low heterogeneity, three moderate, and one very large (*SYNE1*, I^2^ = 79%). Three of the eight regions associated with Wex in the multi-ancestry analysis replicate previously reported signals, while the other five are previously unreported: *1q12.13, FSHR:1, 4q35.2, SCNN1A*, and *TCEA2*. All six regions associated with the narrow phenotypes are previously unreported (Figure 1c). Two of these regions were driven uniquely by non-EUR studies, *1q42.13* (Wex, AFR) and *21q22.13* (PCN.v1/SCN.v1, AMR). The surgically-confirmed phenotype associations displayed the most extreme effect sizes (Figure 1d), including *21q22.13* (rs186666195:T OR_SCN.v1_ = 5.23 and OR_PCN.v1_ = 3.65) and *19q13.42* (rs73061950:A OR_PCN.v1_ = 3.68). Six of the 58 regions from all 11 GWASs had heterogeneity when comparing the 95% odds ratio confidence intervals between the six phenotype definitions. For *MEGF6, 13q22.1*, and *19q12*, the narrow phenotypes had higher effect size estimates than W. For *SYNE1* and *TCEA2*, W had a higher odds ratio than Wex but in contrast, Wex was higher than W for *SCNN1A*. The liability scale heritability captured by the EUR Wex GWAS was 0.097 (0.022). Heritability estimates for the remaining narrow phenotypes were non-significant and are provided in Supplementary Table 8. None of the partitioned heritability estimates for the Wex and narrow phenotypes (EUR-only) reached significance (Supplementary Table 9).

### Fine-Mapping to Detect Causal Variants at GWAS Loci

To characterize shared and population-specific putative causal variants, we conducted statistical fine-mapping (See Methods) on all genome-wide significant regions from multi-population meta-analyses, totaling 63 significant loci representing 57 regions in five of the six studies (the SCN.v2 meta-analysis yielded no genome-wide significant results). Among these, 14 loci contained at least one variant with posterior inclusion probability (PIP) greater than 0.5, corresponding to 37 unique causal variants with strong evidence for causality (Supplementary Table 11). The cross-ancestry fine-mapping analyses computed PIPs for each variant across all combinations of ancestry groups included in the overall study.

Of the 14 regions with at least one variant in the credible set, eight were from the W multi-population meta-analysis, four were from Wex, and there was one each from PCN.v1 and PCN.v2. Twelve of the credible sets included variants detectable in part due to the incorporation of additional genetic ancestry groups (AMR, AFR, and SAS) previously absent from analyses. In particular, three loci (*1p13.2, FSHR:1*, and *LOC730338*) derived their credible sets from incorporating four of the five ancestry groups. All PIPs from MESuSiE are available in Supplementary Table 11.

### GWAS Variant-Set Gene Enrichment Analyses

From 11 meta-analyses, we identified 79 associations with 47 unique gene regions that were enriched for significant GWAS variants. Thirteen associations, representing 11 unique genes, from MAGMA did not overlap with significant GWAS regions, of which 11 were detected by W analyses. While these genes are not in the GWAS catalog, several of them have been implicated via other types of studies in fertility issues^40–42^, endometrial cancer^43^, adenomyosis^44^, and even endometriosis^45,46^. Two genes were significant in both the META and EUR-wide phenotype GWASs: *CCHCR1* and *KCTD13*. The remaining non-W, non-GWAS significant MAGMA hits were for the Wex (*GLB1L3*, P = 3.40 × 10^−7^) and PCN.v1 (*LRRC2*, P = 2.09 × 10^−7^) phenotypes. All significant MAGMA results are reported in Supplementary Table 12.

### Exploring Cross-Trait Pleiotropy Through Genetic Correlation and Mendelian Randomization

Out of 3,065 single-ancestry genetic correlation tests spanning complex trait GWASs from the UK Biobank (UKBB) phenome, the All of Us (AOU) phenome, microbiome, and partial proteome datasets, 891 traits were nominated for bidirectional Mendelian randomization analysis. A total of 135 phenotypes showed significant genetic correlation with endometriosis at a Bonferroni-corrected threshold of P < 1.63 × 10^−5^, including age at first live birth (rg = -0.24), sleeplessness/insomnia (rg = 0.19), headache (rg = 0.30), and antidepressants prescription (rg = 0.39). Evidence for vertical pleiotropy was observed in 94 exposure-outcome pairs, including associations suggestive of endometriosis contributing to risk of hyperlipidemia, prescription of antacids, and reduced levels of Apolipoprotein F and *Bifidobacteriaceae*. Of the 94 putative causal relationships, 15 showed low heterogeneity (I^2^ < 25%) in their Egger test, and two had only moderate heterogeneity (25% < I^2^ < 50%). Nominated genetic correlation traits and their MR results are shown in Figure 2. Detailed results of the analyses are provided in Supplementary Tables 13 and 14 for genetic correlation and MR, respectively.

**Figure 2:**
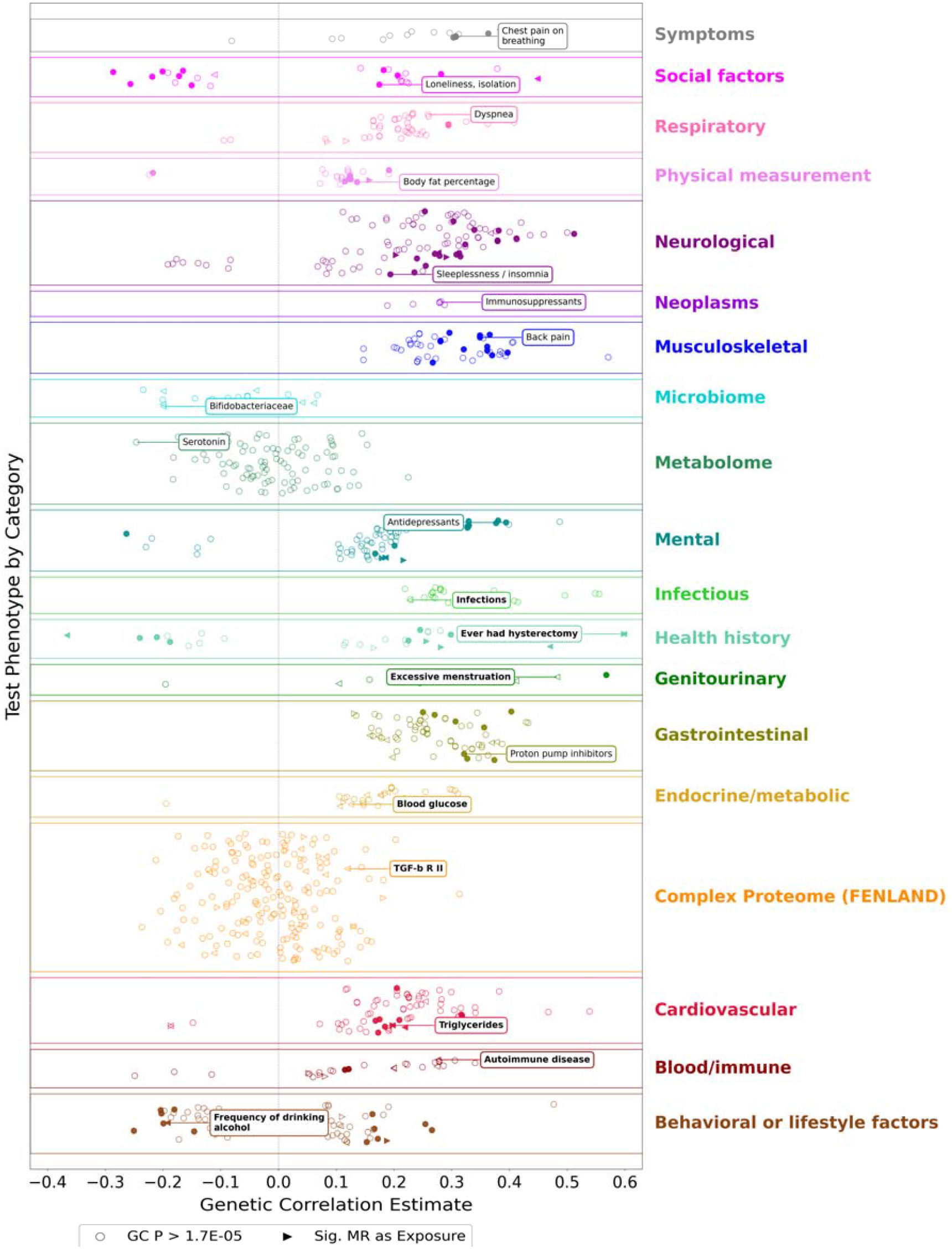
Genetic correlation (GC) and Mendelian randomization (MR) analyses. Each scatter point, colored by category, represents a GC estimate for which the p-value was less than 0.05. Filled shapes indicate a Bonferroni-significant GC value. Triangles indicate significant putative causal relationships detected by MR. One interesting result per category is highlighted with a box. A color fill corresponds to a significant GC estimate whereas a colored outline did not reach multiple testing GC significance. Bold text in the annotation boxes indicates a significant MR pair.

### Identifying Disease-Relevant Cell Types and Crucial Genes

We prioritized 34 cellular contexts (cell type – menstrual group pairs) with significant regression coefficients (P < 2.08 × 10^−4^) for comparison of gene expression among the top MAGMA-identified genes (Figure 3). Regression results of endometriosis status on single cell disease risk score (scDRS) value across all tested cell type-menstrual group pairs are provided in Supplementary Table 15. The smallest cellular context had 22 cells while the largest had 43,999 cells, yielding a wide range of statistical power.

**Figure 3:**
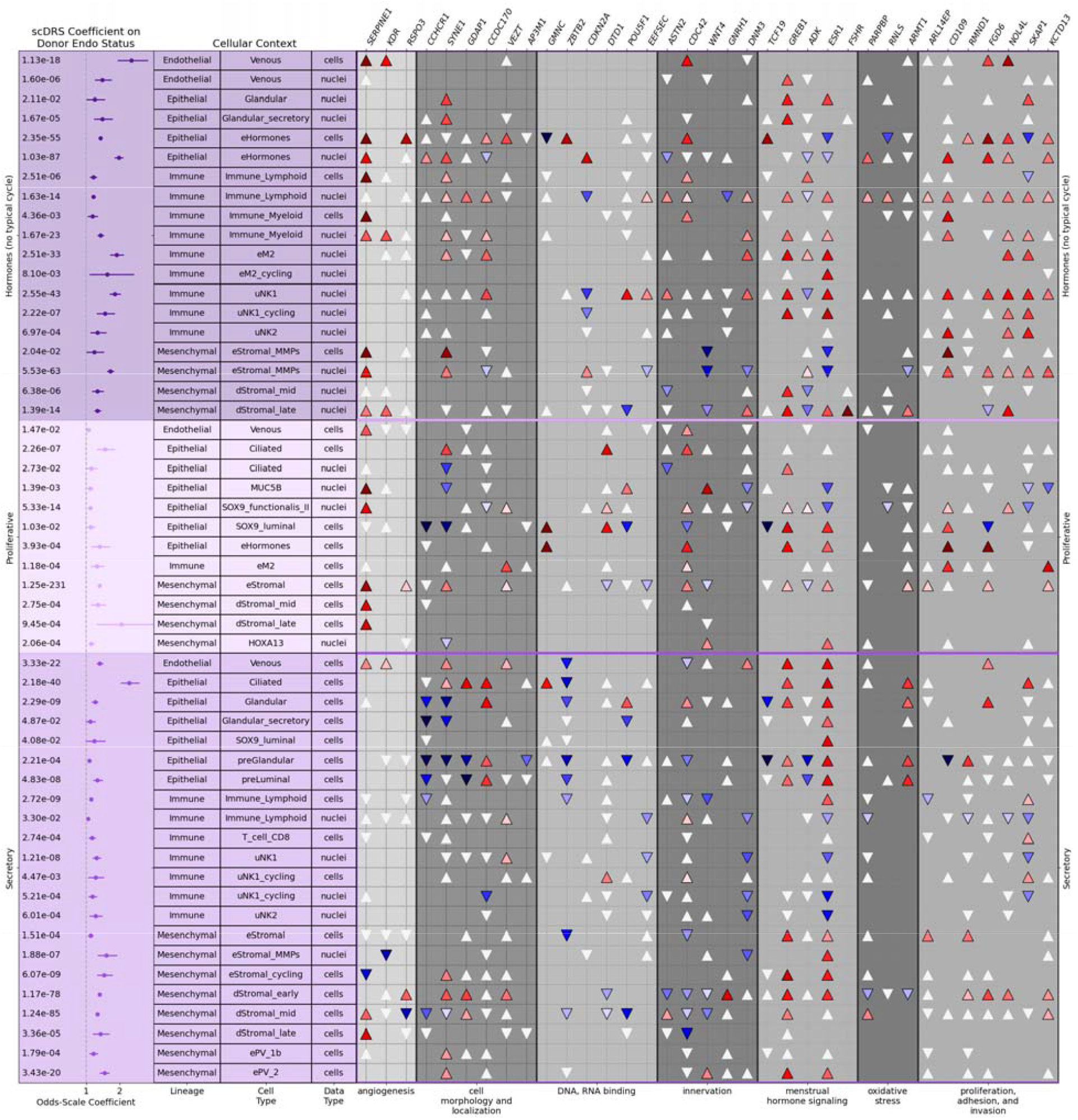
Single cell analyses – disease relevance scores (scDRSs) and differential gene expression. The left purple-colored panels show the odds-scale regression coefficients when comparing cells from donors with endometriosis to donors without endometriosis from different menstrual groups. Each row is annotated with the cell lineage, cell type, and data type. The rows are ordered by cell type. The right gray panels show the results of two-sided t-tests comparing the gene expression of the top MAGMA genes, with significant tests represented by upward tringles for overexpression and downward triangles for underexpression. Uncolored triangles are nominally significant at P < 0.05.

Of the 34 cell contexts with significant regression effect estimates, 14 were derived from donors in the “hormones” menstrual group, five from the proliferative phase, and 15 from the secretory phase. None from the menstrual phase was significant. Of the 207 non-significant contexts, only six had >80% power to detect small effects (log-odds < 0.1) at the Bonferroni-corrected alpha threshold, providing evidence that those effects may be minimal, and conversely suggesting that there may remain underlying effects which were under-powered to detect in the HECA dataset. These contexts spanned all four cellular lineage groups: three endothelial, eight epithelial, ten immune, and 13 mesenchymal. The largest regression coefficient was observed in venous cells from donors on hormones (odds-scale coefficient = 2.34, P = 1.13 × 10^−18^). The most statistically robust signal emerged in endometrial stromal cells sampled during the proliferative phase (odds-scale coefficient = 1.40, P = 1.25 × 10^−231^). Among the genes driving the stratification by each regression model, *ESR1* was most frequently differentially expressed, reaching significance in 23 of the cellular contexts, with 18 of those being overexpression and five demonstrating depletion. *GREB1* was the most consistently upregulated gene (20 contexts: nine hormones, nine secretory, and two proliferative), whereas *ADK* and *ZBTB2* were the most consistently downregulated (7 contexts each). Lymphoid immune nuclei from donors in the hormones menstrual group exhibited the largest number of differentially expressed genes overall (20 genes: 13 upregulated and 8 downregulated). Gene expression results across all cell type–menstrual group contexts are provided in Supplementary Table 16.

Five differentially-expressed genes overlap with four of GWAS regions that had not been previously associated with endometriosis. *FSHR* demonstrated strong overexpression late decidualized stromal nuclei from donors on hormones (Odds-Scale B = 11.0, P = 4.89 × 10^−7^). Two epithelial cell types showed opposing expression effects of *GMNC*, ciliated during the secretory phase (Odds-Scale B = 2.59, P = 1.55 × 10^−6^) and the HECA-defined “endometrial hormones” type^47^ from donors on exogenous hormones (Odds-Scale B = 0.095, P = 2.17 × 10^−60^). *ADK* expression was associated with endometriosis in 11 contexts: three cellular and eight nuclear. Of those 11 contexts, the effect was in the negative direction for seven and positive for four, without a clear pattern. *DTD1* was down-regulated three stromal cellular contexts: non-decidualized (eStromal) during the secretory phase and both early- and mid-proliferative stage.

### Post-GWAS Multi-Omic Analyses Fuel Pathway Identification

To identify associations between endometriosis and additional layers (x) of the central dogma and beyond (p - proteome, s - splice-ome, e - transcriptome, m - methylome, and ed - RNA edit-ome), we performed “x”-ome-wide imputed association studies (xWAS) and xQTL-wide summary-based Mendelian randomization (xSMR), inputting the eleven GWASs’ summary statistics (See Methods). In total, xWAS implicated 57 unique molecular phenotypes, including 2 proteins, 81 splicing junctions, and 36 transcripts (Supplementary Table 17). Putative causal relationships involving 267 unique genes were identified by xSMR, represented by 6 proteins, 79 splicing junctions, 34 eQTLs, 392 CpG sites, and 3 RNA editing A-to-I sites (Figure 4; Supplementary Table 18).

**Figure 4:**
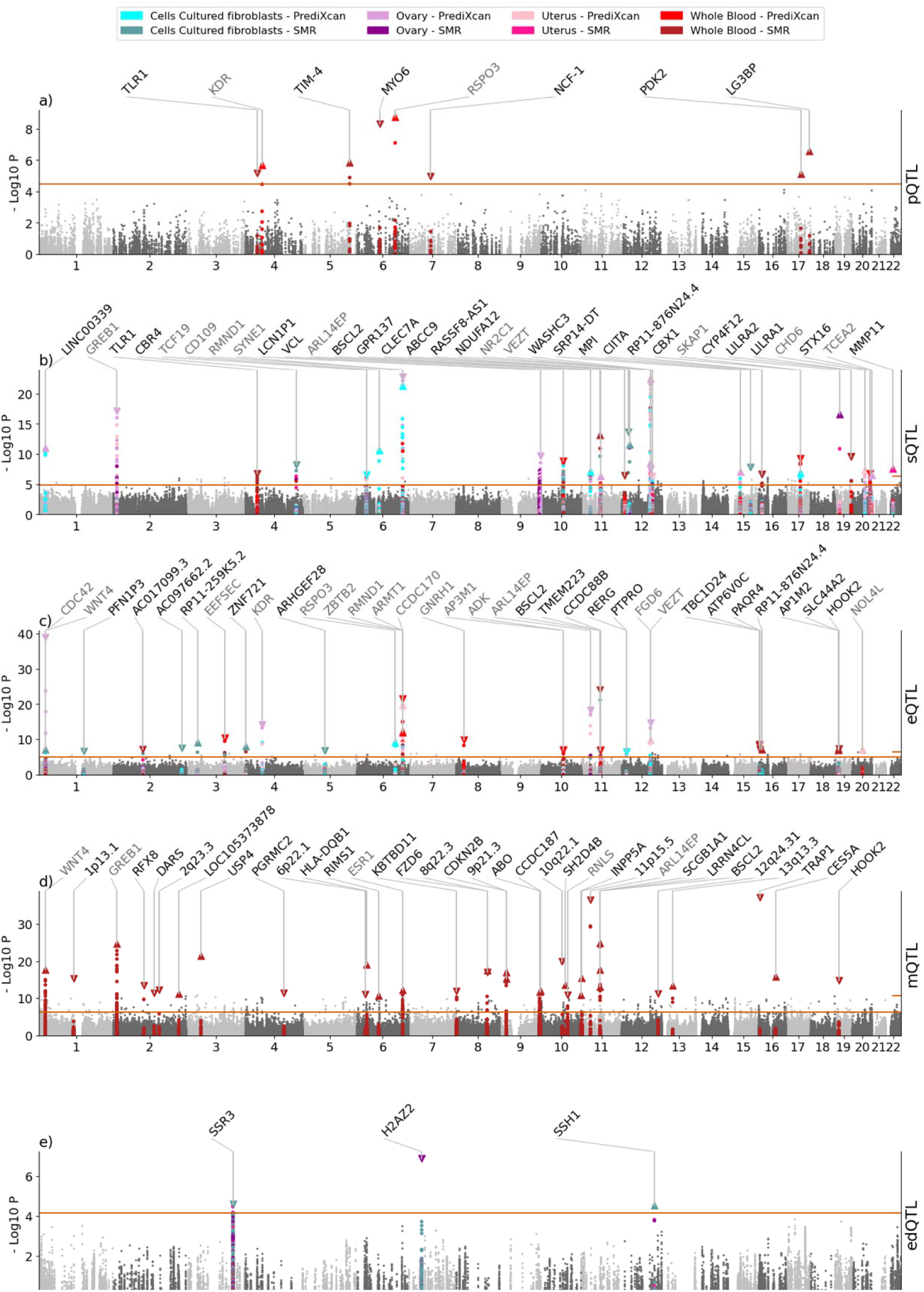
xQTL summary-based Mendelian randomization (xSMR) and PrediXcan imputed “x”ome-wide association study (xWAS) results across four tissue types and five -omic layers: a) proteome, b) splice-ome, c) transcriptome, d) methylome, and e) RNA edit-ome. The p-value thresholds were adjusted for the numbers of tests per analysis (count of sites per combination of GWAS – QTL dataset). The number of genes displayed on each panel is limited to the 35 most significant. Genes annotated in gray overlap with MAGMA genes whereas genes annotated in black do not. Triangles point upward for a positive effect and down for a negative effect.

Of the 299 genes implicated by xQTL summary-based Mendelian randomization (xSMR) and PrediXcan imputed “x”ome-wide association studies (xWAS), 34 overlapped with genome-level associations identified by MAGMA. Above and beyond the GWAS-related genes, pSMR and PWAS detected six proteins, sSMR and SWAS identified introns at 41 genes, eSMR and TWAS revealed 44 genes, mSMR identified methylation sites at 185 genes, and edSMR implicated RNA editing sites at three genes. There were seven previously unreported genes supported by more than two –omics types (including MAGMA), shown in Table 2.

**Table 2:** Previously unreported genes with multiple layers of multi-omic evidence. The minimum GWAS P-value presented is the lowest P-value associated with any of the QTLs tested by xWAS or xSMR. Up arrows indicate positive associations with endometriosis while down arrows represent negative associations with endometriosis. For the transcriptome, the measured quantity is gene expression. For spliceome, it is the excision rate of that junction, and for methylome, it is the rate of methylation at that CpG site.

| Gene | Min GWAS P | MAGMA | Transcriptome | Spliceome | Methylome |
| --- | --- | --- | --- | --- | --- |
| *BSCL2* | W SAS: P = $2.42 \times 10^{-2}$ | N / A | ↓ Fibroblasts<br>↓ Whole Blood | ↓ intron 1, transcript variant 2 or 3 [Fibroblasts, Whole Blood]<br>↑ intron 1, transcript variant 1 [Whole Blood] | ↑ cg15538427 (downstream)<br>↑ cg22231483 (downstream)<br>↑ cg25015585 (downstream)<br>↑ cg23916044 (intergenic, exon 2)<br>↓ cg10573476 (intergenic, intron 1) |
| *CHD6* | W EUR: P = $3.76 \times 10^{-8}$ | W EUR: P = $9.15 \times 10^{-8}$ | ↑ Ovary<br>↑ Uterus | ↓ intron 3 [Fibroblasts, Ovary, Uterus]<br>↓ intron 1 [Ovary, Uterus] | N / A |
| *TCF19* | W META: P = $2.96 \times 10^{-7}$ | W META: P = $1.35 \times 10^{-6}$ | N / A | ↓ intron 3 [Fibroblasts] | ↑ cg22692488 (intergenic, exon 3)<br>↓ cg23819653 (intergenic, exon 4) |
| *AP3M1* | W META: P = $3.08 \times 10^{-8}$ | W META: P = $1.35 \times 10^{-6}$ | ↓ Whole Blood | N / A | ↓ cg26469011 (intergenic, intron 6) |
| *ADK* | W META: P = $1.57 \times 10^{-9}$ | W META: P = $1.98 \times 10^{-11}$ | ↓ Whole Blood | N / A | ↓ cg13085507 (intergenic, intron 1)<br>↑ cg09299075 (intergenic, exon 5) |
| *KCTD13* | W META: P = $1.34 \times 10^{-6}$ | W EUR: P = $2.04 \times 10^{-6}$<br>W META: P = $1.22 \times 10^{-6}$ | ↑ Fibroblasts<br>↑ Ovary | ↓ alternative intron 1 [Whole Blood] | N / A |
| *TCEA2* | Wex META: P = $9.50 \times 10^{-9}$ | Wex META: P = $4.39 \times 10^{-7}$ | N / A | ↓ intron 2, transcript variant 2 [Uterus]<br>↓ skip exon 3 [Ovary, Uterus] | ↓ cg09990613 (upstream)<br>↓ cg12176783 (upstream)<br>↓ cg07774293 (upstream) |

The only pSMR signal which was replicated across multiple case/control definitions was the protein TIM-4, with a positive association across three GWASs: AMR W (B = 0.594, P_SMR_ = 3.26 × 10^−5^), META PCN.v1 (B = 0.402, P_SMR_ = 1.48 × 10^−6^), and META PCN.v2 (B = 0.384, P_SMR_ = 1.27 × 10^−5^). *VCL* was one of seven genes in the intersection of the sSMR with SWAS results, represented by three distinct splicing events with associations across all four tissues, chr10:74109156-74111909 (B = -0.0803, P_SMR_ = 2.81 × 10^−7^, P_SWAS_ = 1.32 × 10^−9^), chr10:74109156-74114184 (B = 0.0692, P_SMR_ = 8.37 × 10^−8^, P_SWAS_ = 2.93 × 10^−9^), and chr10:74112112-74114184 (B = -0.0735, P_SMR_ = 1.51 × 10^−6^, P_SWAS_= 4.52 × 10^−8^). The most robust transcriptome result, with eight associations across both methods, was reduced *EEFSEC* expression, in both whole blood and cultured fibroblast cells (min P = 5.51 × 10^−7^ and min P_TWAS_ = 4.31 × 10^−11^). The gene with the highest number of significant CpG sites (12) from mSMR was *ESR1*. Methylation sites from intron 1 had positive SMR coefficients (highest B = 0.197, P_SMR_ = 2.16 × 10^−7^; probe cg04211581) whereas methylation sites from intron 2 had negative SMR coefficients (lowest B = -0.283, P_SMR_ = 3.02 × 10^−7^; probe cg24764793). Of the three significant RNA-editing sites, the most robust site (chr3:156540497) showed six significant edSMR associations across three tissues. This site in the *SSR3* gene demonstrated stronger effect sizes with the SCN.v1 (surgically confirmed) GWAS than with the wide phenotype (B = -0.0759, P_SMR_ = 2.61 × 10^−5^ for fibroblast cells and B = - 0.0731, P _SMR_ = 3.06 × 10^−5^ for uterus).

Including genomic signals from MAGMA, we identified 314 genes across six -omics types, each supported by respective biological evidence of endometriosis association. We performed chi-squared tests for this gene list with gene sets curated by MSIGDB (Supplementary Table 19) and identified several significant groups. From the Biocarta category of gene sets, there were four sets identified, the CREM (P = 3.83 × 10^−17^), MTA3 (P = 6.06 × 10^−6^), CDC42RAC (P = 2.80 × 10^−5^), and PLC (P = 9.22 × 10^−5^) pathways. Significant regulatory gene sets which had the highest percent coverage in our gene list include target genes of *MIR564* (3 / 5 genes, P = 4.52 × 10^−9^), *MIR6798_5P* (4 / 45 genes, P = 1.25 × 10^−5^), and *MIR11181_5P* (6 / 72 genes, P = 6.14 × 10^−8^). The most significant biological processes (BP) from the gene ontology (GO) with four or more genes overlapping included regulation of integrin activation (P = 6.38 × 10^−7^), mammary gland duct morphogenesis (P = 5.49 × 10^−6^), and lamellipodium assembly (P = 1.43 × 10^−12^). The strongest hit from the REACTOME gene sets was hormone ligand binding receptors (P = 9.07 × 10^−9^).

## Discussion

We report the most diverse of over a million women for GWAS of endometriosis to date incorporating participants from 14 biobanks across the world. Out of 58 associated regions from 11 meta-analyses, 27 are previously unreported while 31 contain known variants. We have specified biological evidence of the genes or proteins relevant to 12 of the previously unreported regions. The associations we detected replicate 37% of the variants in the GWAS catalog, demonstrating consistency with prior findings while also supporting substantial novel discovery in our expanded multi-ancestry, multi-phenotype study. Using a fine-mapping method which leveraged the multi-ancestry nature of our cohorts, we specified 37 putative causal variants. Then, to examine the source of both known and unknown comorbidities, we detected shared genetic architecture between endometriosis and 135 complex polygenic phenotypes. Finally, with the integration of five additional -omics layers, we have implicated a total of 314 genes associated with endometriosis.

Leveraging multi-ancestry data enabled direct comparison of effect sizes across populations and phenotypic definitions. While most regions showed consistent effects, a subset demonstrated ancestry-specific differences, including non-overlapping confidence intervals between ancestries (e.g., *DNM3, 2q35, LOC730338, CCDC53*), suggesting potential population-specific genetic architecture. Similarly, comparisons across phenotype definitions revealed variability in effect sizes at several regions (e.g., *MEGF6, SYNE1*), indicating that genetic effects may differ by disease definition or severity. Overall, heterogeneity was low to moderate for most regions, supporting consistent effects across cohorts, with only two showing substantial heterogeneity. The very large heterogeneity observed at the *SYNE1* region is likely due to multiple independent signals as shown in the conditional analysis, while the large heterogeneity from the *2p13.3* locus comes from a range of effect estimates, all with matching directions. Together, these findings highlight both the overall consistency of genetic risk and the added resolution gained from diverse ancestries and refined phenotyping.

Our ancestry-stratified meta-analyses revealed a consistent observed SNP heritability of 9-13% for endometriosis for four studied genetic ancestry populations with sufficient size (AFR, AMR, EAS, and EUR), which is higher than previously observed SNP heritability of 7% based on a majority-European population^11^. The higher and relatively consistent SNP heritability we observed can be attributed to the unprecedented scale and diversity of our study cohort. For the EAS and EUR W GWASs, the proportions of heritability attributable to enhancers active in uterine tissue (33% and 20%, respectively) were similar to the proportion that of the enhancer annotation in *Finucane et al 2015* (24%)^48^.

The previous endometriosis GWASs have not included any African-ancestry (AFR), Admixed-American-ancestry (AMR), or Central/South Asian (CSA+SAS) populations, which were essential in the computation of 12 / 14 (96%) credible sets in our fine-mapping analysis. We detected three AFR-specific regions (*1q42.13, 2p13.3*, and *20q13.2;* nearest coding regions *ZNF678, CD207*, and TSHZ2) and one AMR-specific region (*21q22.13* : *HLCS*) whose lead variants are rare in other super-populations^49^. We suspect that in addition to the variants being rare, we were limited when identifying genes with *in silico* biological evidence for these regions because the multi-omics datasets we integrated mostly consisted of EUR individuals. The nearest genes specified here were not detected from xWAS or xSMR, and their lead SNPs have no QTLs reported in GTEx. *ZNF678* is most highly expressed in nerve, thyroid, and reproductive tissues^50^. *CD207* is a key player in immune antigen processing^51^, while *TSHZ2* is a tumor suppressor^52^, and *HLCS*, by contrast, may promote breast cancer metastasis^53^. While enabling further investigation into the biological significance of these ancestry-specific associations, we were able to capture a more comprehensive genetic landscape of endometriosis by incorporating biobanks from various regions and ancestry backgrounds worldwide.

Given that diagnosis codes from the EHR can lack specificity for accurately phenotyping endometriosis^54^, we employed narrow phenotyping algorithms which incorporated procedure codes (surgical and imaging). While the W phenotype only yielded an observed prevalence of 4.73%, the most specific phenotypes (confirmed cases and controls) helped bring the ratio of cases and controls closer to the previously-estimated population prevalence of 10%. We also saw potential evidence for ongoing diagnostic bias in that EUR had the highest prevalence for the wide phenotypes, but lowest for PCN.v2 and SCN.v2. This may be indicative of EUR patients being recommended for surgical care at higher rates. Even though the Wex and narrow phenotypes yielded diminished sample sizes, there were still 14 genome-wide significant regions. MAGMA detected regional associations with three additional genes (*LRRC2* with PCN.v1 and *GLB1L3* and *TCEA2* with Wex) that were not detected in the largest GWAS.

Leveraging multi-modal EHR data for more precise phenotyping enhanced our ability to refine and validate genetic associations and uncover deeper biological insights.

Endometriosis has multifaceted risk factors and complex clinical presentations with a wide range of comorbidities and concomitant conditions. We explored genetic correlation and pleiotropy with endometriosis on an unprecedented scale (3,065 tests), leveraging phenome-wide summary statistics from All of Us and the UK Biobank along with metabolite, microbiome, and complex proteome GWASs. With the addition of Mendelian randomization, we have yielded invaluable hypothesis-generating results for further validation, even if genetic variants do not always meet the requisite instrumental variable assumptions for detecting causality. Five metabolites showed nominally significant (p < 0.05) genetic correlation with endometriosis, including serotonin which is known to be related to endometrial stromal cell decidualization^55^ but is also impacted by SSRIs like citalopram which may improve endometriosis symptoms^56^. We further observed evidence of vertical pleiotropy across multiple exposure-outcome pairs, including associations suggestive of hyperlipidemia risk, altered levels of Apolipoprotein F, and shifts in gut microbiome composition, including reduced abundance of *Bifidobacteriaciae* (order *bifidobacteriales*, class *actinobacteria*), which plays a role in estrogen regulation^57,58^. Together, these findings raise the possibility that endometriosis may influence downstream biological pathways relevant to long-term conditions, including cardiometabolic disease, potentially acting as a mediator through immune, metabolic, and microbiome-related mechanisms. While these observations require further validation, they provide a framework for understanding how endometriosis may contribute to broader systemic disease risk beyond reproductive health.

One hallmark of endometriotic lesions is cellular adhesion, the process by which cells migrate and interact with the extracellular matrix^59,60^. An essential cytoskeletal protein which plays a role in regulating adhesion and motility is vinculin (Vcn)^61^. We detected three endometriosis-associated splicing events via sSMR and SWAS which support that expression of an isoform of Vcn, metavinculin (MVcn, +exon 19), may reduce endometriosis risk. The AA genotype at rs7896966 is associated with increased inclusion of exon 19^62^, and our GWAS identified that the AA genotype is also associated with decreased rates of endometriosis. It has been demonstrated that MVcn expression contributes to fewer but larger focal adhesions per cell^61^ and that the inhibition of focal adhesion via FAK decreases the formation of endometriotic lesions in mice^60^. While the GO gene set for focal adhesion assembly was not statistically significant, there were four genes that overlapped with that pathway: *VCL* (sSMR), *WNT4* (mSMR, uterus- and ovary-TWAS), *ARHGEF7* (mSMR), and *KDR* (six cellular contexts, mSMR, PWAS, and fibroblast-, uterus-, and ovary-TWAS). *RHOD* expression (codes for the GTPase RhoD) had a negative causal relationship with endometriosis, which aligns with previously published results that show that silencing RhoD leads to less efficient cell migration^63^. *KDR* exhibited mixed effect directions. Plasma PWAS along with endothelial cells and decidualized stromal cells supported positive associations whereas TWAS and stromal matrix metalloproteinase cells from the secretory phase indicated a negative association with *KDR* expression. When KDR (a.k.a. vascular endothelial growth factor receptor 2; *VEGFR2*) is stimulated by vascular endothelial growth factor (VEGF), it stimulates DNA synthesis and promotes focal-adhesion-related cell migration^64^. Although more investigation is required to precisely elucidate how the proteins in this pathway interact in the context of endometriosis, our data clearly support the hypothesis that the regulation of focal adhesion may be a strong candidate for therapeutic targets^60^.

Given the strong interplay between endometriosis and regulation of the menstrual cycle, it is important to take note of genetic and molecular signatures which may indicate hormonal dysregulation. *GREB1* (codes for Growth Regulating Estrogen Receptor Binding 1) is a well-characterized endometriosis-associated gene. We identified 20 cellular contexts in which *GREB1* expression is elevated in cells from donors with endometriosis, mostly from the cells and nuclei sampled during the secretory phase and from donors exposed to exogenous hormones. All five associations between elevated *GREB1* and endometriosis under the immune lineage were in nucleus types from donors on hormones. Since hormones are a common first-line treatment of endometriosis symptoms^65^, it is important to investigate how these medications interact with immune cell types that contribute to an inflammatory response. We have replicated associations with known methylation and splice sites at the *GREB1* locus^66^, while also expanding upon the multi-omic molecular signature. *GREB1* has a truncated isoform, GREB1c, and we demonstrate that splicing events in ovarian and uterine tissue associated with this isoform increase risk of endometriosis while splicing events associated with the canonical isoform (GREB1a) are associated with decreased disease risk. Importantly, *GREB1* was not associated with endometriosis on the eQTL or pQTL levels, emphasizing that in order to promote mechanistic hypotheses of endometriosis, we must continue to incorporate comprehensive layers of multi-omics.

Despite the large sample size, heritability observed with GWAS (10-13%) still fails to measure up to the broad sense heritability estimation from a twin study of 47%^14^. Rare variants, structural variants, nonlinear effects, or gene-environment interactions might contribute to endometriosis risk but remain undetected in our analyses. As we leveraged the GWAS results to study other -omics from the central dogma and beyond, one limitation was that we lacked individual-level data. This means we relied on the thousand genomes dataset as an LD reference panel, which has limited size and may create LD mismatch with the study population. We suspect that is why we only were able to detect putative causal variants in 14 / 57 regions. The results captured by xWAS and xSMR relied on previously-trained *cis*-xQTL models that are limited to capturing only genotype-driven molecular trait variability. This may leave out complex genetic architecture (*i.e. trans*-xQTLs) or environmentally-driven molecular signatures potentially relevant to our phenotype of interest^67^. While our comprehensive *in silico* multi-omic approach yielded many testable hypotheses, we did not perform any experimental functional validation. Furthermore, although our GWAS constitutes the most diverse genetic study of endometriosis performed so far (32% non-European), we still lacked the power to draw many meaningful conclusions in understudied populations. We want to emphasize that the field should continue prioritizing the inclusion of diverse cohorts in genetic studies.

In addition to replicating findings from previous studies, we uncovered new insights that advanced our understanding of the genetic underpinnings of endometriosis. Our integrative multi-omics approach, combining genomic data with single-cell analyses along with proteomic, splice-omic, transcriptomic, methylomic, and RNA edit-omic tests, has biologically implicated 314 genes, providing unprecedented insights into the molecular mechanisms driving this chronic condition. These findings lay a robust foundation for future functional studies to elucidate the precise roles of identified genes and pathways in endometriosis pathogenesis.

Moreover, our results have important clinical implications, potentially informing the development of more accurate diagnostic tools, personalized risk prediction models, and targeted therapeutic interventions. As we move forward, this work emphasizes the critical importance of large-scale, diverse genetic studies in unraveling the complexities of multifactorial diseases like endometriosis, paving the way for improved patient outcomes and a deeper understanding of women’s health issues globally.

## Methods

### Phenotyping

Endometriosis diagnosis requires direct observation of lesions by laparoscopic surgery or sometimes imaging (MRI/ultrasound). We can extract diagnoses from medical records via diagnosis billing codes in the form of ontologies like the international classification of diseases (ICD) or the systematized nomenclature of medicine (SNOMED). For the EHR-linked biobanks, we used structured data to define the phenotypes. Cases for wide endometriosis (W) were any women with a history of ICD-9 (617.*), ICD-10 (N80.*), or SNOMED codes for endometriosis. For the wide phenotype excluding adenomyosis (Wex), women with a history of ICD-9 (617.0), ICD-10 (N80.0), or SNOMED codes for uterine endometriosis were excluded from both cases and controls.

Our two narrow case definitions were procedure-confirmed and surgically-confirmed (PCN and SCN). These narrow phenotypes were designed to capture confirmed cases and controls of endometriosis based on procedure history including hysterectomies, laparoscopies and ultrasounds. The list of procedure codes (CPT-4 or OPCS) for PCN included all the surgery codes in SCN but added non-obstetric ultrasound codes to account for potential imaging diagnoses. Each of the two narrow phenotypes was tested in two versions: cases versus all controls (PCN.v1 and SCN.v1) and cases versus confirmed controls (PCN.v2 and SCN.v2) where confirmed controls were those who had history of the corresponding procedure codes for each phenotype with no history of endometriosis diagnosis. For additional details on the phenotyping algorithms, see Supplemental Appendix 1. To compare case proportions across ancestry groups for each of the phenotypes, we performed pairwise two-sided Z-tests. Significant Z-tests were those with P < 1.32 × 10^−3^ (0.05 / 38 comparisons).

### Genotyping by Biobank

Most contributing biobanks genotyped several hundred thousand variants. After those data are collected, then the rest of the genomic variants can be imputed probabilistically based on a reference panel such as TOPMED. Some biobanks such as AOU used short read whole genome sequencing (WGS) to gather their genomic data. The sequences from the short reads are aligned and compared to the reference to produce variant calls. References and platform information for each biobank’s genotyping method and QC can be found in Supplementary Table 20.

### Genetic Association Testing

Each contributing biobank performed ancestry-stratified association testing with up to six phenotypes (W, Wex, PCN.v1, PCN.v2, SCN.v1, and SCN.v2), depending on whether they had at least 50 cases for that phenotype and ancestry combination. Following with prior protocols from GWAS meta-analysis consortia^15,17,68,69^, we set this lower threshold for the number of cases to prioritize including underrepresented ancestry groups. Genetically-informed ancestry (GIA) is defined based on genetic distance from subpopulations of a reference group such as the 1000 Genomes Project dataset^70^ (1KG). Genetic distance is measured using a dimensionality reduction technique such as principal component analysis (PCA). Ancestry is then assigned using a classifier such as a mixture model or k-nearest neighbors. For references and methods used for assigning GIA within each biobank, see Supplementary Table 21.

Contributing biobanks used linear mixed models to estimate the effect of each variant on the six phenotypes. Tools implemented for these tests include SAIGE^71^ and Regenie^72^. Association tests were adjusted for principal components, age at EHR data collection, and any biobank-specific batch variables (for example, collection site within eMERGE). For estimation of the null models (SAIGE or Regenie step 1), variants were required to have a call rate of at least 95%, a minimum minor allele frequency of 0.01, and a Hardy-Weinberg p-value of at least 1×10^-6^. For association testing (SAIGE or Regenie step 2), variants were required to have a minor allele count of at least 20, and an imputation quality of at least 0.6. Individuals were included if their overall genotyping rate was at least 95%. A summary of each biobank’s exact covariates and QC procedures is in Supplementary Table 22. We leveraged publicly-available FinnGen^73^ summary statistics to increase the sample size.

After biobank analyses were completed and collected, summary statistics were cleaned and lifted into genome build hg38 as necessary. Then, inverse variance-weighted, fixed-effect meta-analyses were performed using GWAMA (software for genome-wide association meta-analysis)^74^. GWAMA adjusts each study based on its overall genomic inflation factor. Prior to meta-analysis, input summary statistics were restricted to variants with a minor allele frequency of at least 0.005 to ensure stability in the estimates. GWAMA also returns Q values, from which we calculated I^2^ percentages. For lead SNPs, we categorized low heterogeneity as 0%-25%, moderate as 25%-50%, high as 50%-75% and very high as 75%+ as defined in *Ioannidis, et al 2007*^*75*^.

To contextualize novel findings relative to prior studies, we assessed replication among individual variant associations by examining the effect sizes and p-values from the 11 GWASs for 177 unique genome-wide significant endometriosis-associated variants from the GWAS catalog,^37^ in addition to 16 hits from a Taiwanese-Han GWAS,^76^ which are not available in the GWAS catalog. Then, to define significantly-associated loci, we started by applying the clumping function of Plink 1.9^77^ with the lead SNP p-value threshold set to 5×10^-8^, the secondary SNP threshold set to 1, minimum R^2^ of 0.1, and a window of 1000kb. 1KG was used as a linkage disequilibrium (LD) reference, matching the whole dataset to the multi-ancestry meta-analyses and single-ancestry subsets of the dataset to their respective meta-analyses. After the variant clumps were identified, any clumps that were physically overlapping with one another were merged into loci. For each locus, we performed summary statistics-based conditioning analysis with GCTA-COJO^78^. 1KG was used for LD reference, and the default collinearity threshold of 0.9 was used.

To describe study-wide associations, we merged overlapping per-GWAS loci into “regions.” Then, we tested if each region contained any endometriosis-associated variants reported in the GWAS catalog^37^, searching for the terms “endometriosis” and EFO:0001065^21^. Each locus is labeled with its cytoband, but if the corresponding gene region was statistically significant in the MAGMA, xSMR, or xWAS tests (described below), or if the GWAS locus is in contact with a gene based on endometrial cell chromatin looping data^79^, that gene symbol was added to the locus label.

### Estimation of Observed SNP Heritability and Genetic Correlation Amongst Endometriosis GWASs

We estimated heritability with the LD-Score Regression tool, LDSC^80^, for all single-ancestry meta-analyses representing at least two thousand cases. LD scores were computed for HapMap variants^81^ from the respective super-population subgroups from 1KG for each ancestry-stratified meta-analysis. We also estimated partitioned heritability^48^ with variant annotation groups coming from ENSEMBL regulatory features GRCh38.105^82^ and uterus-specific regulatory features from EpiMap^83^. We estimated cross-ancestry genetic correlation of our meta-analyses using Popcorn^38^. Popcorn cross-population LD scores were derived from the same set of variants and samples from 1KG as in LD score computation for LD score regression.

### Statistical Fine-Mapping for Identification of Putative Causal Variants

The multi-ancestry sum of the single effects model (MESuSiE)^84^ was used to identify putative causal variants in each significant GWAS locus from the multi-ancestry meta-analyses. For each locus, we extracted the single-ancestry summary statistics from that region in addition to computing ancestry-specific LD matrices for AFR, AMR, EAS, EUR, and SAS. MESuSiE analyzes whether causal variants are shared between ancestry groups by computing a posterior inclusion probability (PIP) for each ancestry alone and for each combination of two or more ancestries. The maximum number of effects to consider (L) was set to 10 by default, and SNPs with any PIP value greater than 0.5 are part of the credible set for that locus^84^.

### Variant-Set Enrichment for Identifying Enriched Gene Regions

MAGMA^85^ was used to assess gene regions within the GWAS results for enrichment. MAGMA uses a model based on multiple regression to test the association of a phenotype with groups of variants. Testing groups of variants increases statistical power over the single-variant GWAS tests performed^85^. We utilized gene region definitions from NCBI for human genome build 38, testing between 18,691-18,846 genes for each GWAS meta-analysis (MAGMA test counts and p-value thresholds are available in Supplementary Table 23).

### Phenome-Wide and Multi-Omic Genetic Correlation and Mendelian Randomization Tests

The AFR, EAS, and EUR W GWASs were utilized for LDSC^80^ genetic correlation (GC) tests with phenome-wide GWAS summary statistics from AOU^86^ (AFR, EAS, and EUR) and UKB (EUR-only). We only estimated genetic correlation for GWAS pairs with matching, single-ancestry groups. In AOU, we tested for correlation with binary traits that had at least 2,000 cases (medications and phecodeX phenotypes) and quantitative traits with at least 2,000 samples overall (labs and biometric measurements), totaling 1,728 tests. From UKB, we utilized summary statistics for diagnoses and survey variables, both binary and quantitative, with heritability estimates for which the Z-score was at least 4 (527 studies)^87^. European-ancestry summary statistics were also obtained for 169 metabolites^88^, 165 gut microbiome taxa^89^, and 487 proteins^90^ which we were unable to map to one genomic location (mappable proteins were utilized in SMR described below). Reference LD scores were computed from the corresponding 1KG superpopulation groups across Hapmap variants^81^. For the larger-sample tests (AOU and UKBB phenome), genetic correlation results with a p-value of < 0.05 were candidates for mendelian randomization, and for the smaller tests (proteome and microbiome), traits with an h^2^ p-value or rg p-value < 0.05 were nominated. Effect estimates were not available for the metabolites, so we could not perform MR on those. Significant GC estimates were those that reached a p-value < 1.63 × 10^−5^ (correcting for 3,065 total tests).

To distinguish horizontal and vertical pleiotropy, we performed secondary mendelian randomization (MR) tests on the 891 nominated traits using the MendelianRandomization R package^91^. Several MR methods were utilized as a sensitivity analysis for the estimates: inverse-variance weighted (IVW), debiased IVW, lasso-penalized IVW, weighted median, maximum likelihood, and MR-Egger (random-effects). We tested in both directions: endometriosis causing the test trait and vice-versa). Instrumental variables (IVs) were the genome-wide significant variants from the HapMap-filtered^81^ exposure summary statistics, and we used correlation matrices calculated with PLINK 2.0^92^ to adjust for the covariance between IVs. Significant causal relationships were those for which the maximum p-value across methods was < 0.05, the minimum p-value was < 2.84 × 10^−5^ (1,760 total bidirectional tests), all estimates were the same direction, and that direction matched the direction of the genetic correlation estimate. When displaying GC and MR results, we selected pairs that were among the strongest effect sizes in their categories which also relate to known comorbidities of endometriosis.

### Integration with Single-Cell Data

Single cell data are rich resources for understanding the biology of diseases. A recently published endometrial single cell atlas (HECA) includes gene expression data for over 600,000 cells, nuclei, immune cells, and immune nuclei, covering over 40 different cell types, four menstrual groups (three phases and no cycle due to exogenous hormones), and four different endometrial pathologies. The scRNA-seq and snRNA-seq data from 63 donors in four datasets were merged and harmonized before QC and cell type assignment according to gene expression. The data are publicly available as normalized values in ScanPy format^47,93^. We leveraged these data to compute single-cell disease-relevance scores (scDRSs) based on the associated genes from GWAS, as computed by MAGMA. Disease risk scores were computed and normalized using the scDRS software^94^.

For each cell type and menstrual group (“cellular context”), we computed the logistic regression coefficient of the scDRS comparing cells from donors with and without endometriosis. These regression models were adjusted for age; one source dataset only had an age range (18-36), so we assigned age with a random uniform distribution to each of those cells. These regression coefficients were used to prioritize cellular contexts (cell type – menstrual group pairs). Significant contexts were those which had a coefficient one-sided p-value of < 0.05 / 240 tests. For prioritized contexts, we compared the expression of the top MAGMA genes (36 genes) between case and control donors using another set of age-adjusted logistic regressions to identify which genes were driving the distinguishing effect of the scDRS. Significant coefficients were those with a p-value < 0.05 / 1,224 (36 genes x 34 significant contexts).

### Multi-Omic Imputed Association Studies and Mendelian Randomization

We used GWAS summary statistics from the 11 meta-analyses to estimate the association between endometriosis, transcript expression (TWAS), protein expression (PWAS), and intron inclusion (SWAS). For TWAS and SWAS, we used precomputed GTEx v8 eQTL and sQTL weights, respectively, from the PredictDB data repository (http://predictdb.org/) which represented *cis*-level associations in uterine tissue, ovarian tissue, whole blood, and cultured fibroblast cells. Uterine and ovarian tissue were selected because of their relevance to hormone dysregulation, whole blood was included to detect systemic markers, and fibroblast cells were included because endometriotic lesions are fibrotic. For PWAS, we used whole blood pQTL models from *Schubert et al*^*95*^ which are based on the SOMAscan assay data measured in multi-ethnic Trans-omics for Precision Medicine (TOPMed) Multi-omics pilot study. We used baseline models only (no covariates) which had a predictive correlation (rho) of at least 0.1 and a z-score p-value threshold of 0.05, ensuring that the association between the SNPs and protein expression was statistically significant. The xQTL models along with the GWAS summary statistics were the input for summary-based PrediXcan (S-PrediXcan)^96^. We used Bonferroni-corrected p-value thresholds to identify statistically significant associations for each tissue study. The alpha value was 0.05 and the numbers of tests for each GWAS-tissue-QTL combination can be found in Supplementary Table 24. For labeling sQTLs, GTEx provided a mapping from introns to transcripts.

Summary-Based Mendelian Randomization (SMR) is a publicly-available softare tool which leverages uploaded GWAS summary statistics to detect pleiotropic associations between molecular phenotypes as the exposure and a trait of interest as the outcome^97^. We tested all combinations between the 11 GWASs and the following QTL datasets: GTEx eQTLs^98^, sQTLs^98^, and RNA editing QTLs (edQTLs)^99^ for ovary, uterus, whole blood, and fibroblast cells, plasma pQTLs from the FENLAND study^90^, McRae whole blood mQTLs^100^, and Hatton blood mQTLs derived from EAS and EUR populations^101^. We included edQTLs because they show enriched heritability for inflammatory conditions^99^. Base modification from adenosine to inosine can be identified in RNA-seq, and the ratios of modification at those sites were used in QTL analysis^99^. The quantitative molecular traits were the exposures with endometriosis being the outcome. Significant causal associations were identified as those with a p-value less than 0.05 / the number of tests for each GWAS-tissue-dataset combination. Then, significantly heterogeneous results (HEIDI P < 0.05 / the number of significant tests) were discarded; molecular traits with fewer than three instrumental variants were not tested for heterogeneity. The test counts and the corrected p-value thresholds for the xSMR analyses are available in Supplemental Table S25.

Once we aggregated all genes resulting from MAGMA, xWAS, and xSMR, we performed basic gene set testing using chi-squared statistics comparing our gene list to each pathway’s list of genes. The gene sets were from MSIGDB and included hallmark pathways, cell type signatures, co-regulation pathways, and GO ontology pathways. The numbers of gene set tests corrected for are available in Supplementary Table S26.

## Supporting information

All Supplemental Tables

## Data Availability

All data produced in the present study are available upon reasonable request to the authors.

## Data and Code Availability

All data required to recreate figures are available in supplemental tables S1-S26. GWAS summary statistics will be uploaded to the GWAS catalog upon manuscript acceptance.

The availability of individual-level data varies by biobank. AOU, eMERGE, and UKBB are available to researchers by application. The other biobanks (BBJ, BioVU, CCPM, FinnGen, GHI, HUNT, Latvian National Biobank, MGBB, MVP, PMBB, and TWB) only allow access to institutional researchers via their respective IRB protocols.

Code written and run by the coordinating center (PMBB) is available in this github repository: https://github.com/Setia-Verma-Lab/guare_et_al_endometriosis_gwas_2025.

## Acknowledgments

Research reported in this publication was supported by the Eunice Kennedy Shriver National Institute of Child Health and Human Development of the National Institutes of Health under award number R01HD110567.

We acknowledge the Penn Medicine BioBank (PMBB) for providing data and thank the patient-participants of Penn Medicine who consented to participate in this research program. We would also like to thank the Penn Medicine BioBank team and Regeneron Genetics Center for providing genetic variant data for analysis. The PMBB is approved under IRB protocol# 813913 and supported by Perelman School of Medicine at University of Pennsylvania, a gift from the Smilow family, and the National Center for Advancing Translational Sciences of the National Institutes of Health under CTSA award number UL1TR001878.

This phase of the eMERGE Network was initiated and funded by the NHGRI through the following grants: U01HG008657 (Group Health Cooperative/University of Washington); U01HG008685 (Brigham and Women’s Hospital); U01HG008672 (Vanderbilt University Medical Center); U01HG008666 (Cincinnati Children’s Hospital Medical Center); U01HG006379 (Mayo Clinic); U01HG008679 (Geisinger Clinic); U01HG008680 (Columbia University Health Sciences); U01HG008684 (Children’s Hospital of Philadelphia); U01HG008673 (Northwestern University); U01HG008701 (Vanderbilt University Medical Center serving as the Coordinating Center); U01HG008676 (Partners Healthcare/Broad Institute); and U01HG008664 (Baylor College of Medicine).

We gratefully acknowledge All of Us participants for their contributions, without whom this research would not have been possible. We also thank the National Institutes of Health’s All of Us Research Program for making available the participant data examined in this study.

This research has been conducted using the Taiwan Biobank Resource. We thank all the participants and investigators of the Taiwan Biobank. We thank the National Center for Genome Medicine, Academia Sinica, Taiwan, for the technical support in the genotyping. We thank National Core Facility for Biopharmaceuticals (NCFB, 112-2740-B-492-001) and National Center for High-performance Computing (NCHC) of National Applied Research Laboratories (NARLabs) of Taiwan for providing computational resources and storage resources. This study was supported by the National Health Research Institutes (NP-109, 110, 111, 112, 113-PP-09), Ministry of Science and Technology (MOST 109-2314-B-400-017, 110-2314-B-400-028-MY3), and National Science and Technology Council (NSCT 113-2628-B-400-002), Taiwan.

The Trøndelag Health Study (The HUNT Study) is a collaboration between HUNT Research Centre (Faculty of Medicine and Health Sciences, NTNU, Norwegian University of Science and Technology), Trøndelag County Council, Central Norway Regional Health Authority, and the Norwegian Institute of Public Health. The genotyping in HUNT was financed by the National Institutes of Health; University of Michigan; the Research Council of Norway; the Liaison Committee for Education, Research and Innovation in Central Norway; and the Joint Research Committee between St Olavs hospital and the Faculty of Medicine and Health Sciences, NTNU. The genetic investigations of the HUNT Study are a collaboration between researchers from the HUNT Center for Molecular and Clinical Epidemiology (formerly known as the K.G. Jebsen Center for Genetic Epidemiology as of August 1st 2023), NTNU, and the University of Michigan Medical School and the University of Michigan School of Public Health. We thank HUNT participants for donating their time, samples, and information to help others; clinicians and other employees at Nord-Trøndelag Hospital Trust for their support and for contributing to data collection.

The authors thank Million Veteran Program (MVP) staff, researchers, and volunteers, who have contributed to MVP, and especially participants who previously served their country in the military and now generously agreed to enroll in the study.

Support for title page creation and format was provided by AuthorArranger, a tool developed at the National Cancer Institute.

