## Supplemental Appendices for "Expanding the genetic landscape of endometriosis: Integrative -omics analyses implicate key genes and pathways in a multi-ancestry study of over one million women"

### Appendix 1: Detailed Phenotyping Methods

Below are the phenotyping instructions provided to each biobank, including lists of diagnosis codes (ICD and SNOMED), lists of procedure codes (CPT4), and an algorithm for deriving procedure-confirmed cases and controls.

#### Diagnosis Codes

##### "Endometriosis"

- ICD-10: N80.*
- ICD-9: 617.*
- SNOMED: 5562006, 22611009, 53913001, 57493005, 76376003, 129103003, 198251001, 266589005

##### "Adenomyosis/Uterine Endometriosis"

- ICD-10: N80.0*
- ICD-9: 617.0*
- SNOMED: 76376003

#### Procedure Codes

Include procedures that take place at age 55 or younger.

##### "Surgery" (CPT4)

49322, 51992, 57425, 58541, 58542, 58543, 58544, 58546, 58548, 58550, 58552, 58553, 58554, 58570, 58571, 58572, 58573, 58660, 58661, 58662, 58670, 58671, 58673, 58679, 58660, 58661, 58662, 58670, 58671, 58673, 58679

##### "Procedure" (CPT4)

All "Surgery" Codes PLUS ultrasound codes: 76830, 76856

#### Procedure-Confirmed Case-Control Algorithm

```
dx_cases = set of IDs with ICD/SNOMED diagnosis
procedure_cases = set of IDs with CPT4 procedure (age < 55)

confirmed_controls = set of IDs with procedure AND NOT dx
ambiguous_cases = set of IDs with dx AND NOT procedure
potential_conf_cases = set of IDs with dx AND procedure

confirmed_cases = empty list

for potential_case in potential_conf_cases:
    dx_dates = dates of encounters with ICD/SNOMED codes
    proc_dates = dates of procedure codes

    # time window = dx_date within 30 days before and
    # 60 days after procedure
    if any dx_dates and proc_dates are within time window:
        add potential_case to confirmed_cases
    else:
        add potential_case to ambiguous_cases
```

| | |
|---|---|
| V1 Analyses: | confirmed_cases |
| | everyone except for ambiguous_cases |
| V2 Analyses: | confirmed_cases |
| | confirmed_controls |

### Summary of 2 Wide and 4 Narrow Case-Control Phenotypes

**<u>All Analyses are Female-Only (Ideally Determined by Genetic Sex)</u>**

- W - Endometriosis
  - Cases = Endometriosis
  - Case Exclusions = None
  - Control Exclusions = None
  - Procedures = None
- Wex - Endometriosis excluding adenomyosis from cases and controls
  - Cases = Endometriosis
  - Case Exclusions = Adenomyosis
  - Control Exclusions = Adenomyosis
  - Procedures = None
- PCN.v1 - Procedure-Confirmed Endometriosis vs All excluding “ambiguous cases”
  - Cases = Confirmed Endometriosis
  - Controls = Non-Cases Except for “Ambiguous Cases”
  - Procedures = Procedure
- PCN.v2 - Procedure-Confirmed Endometriosis vs “confirmed controls”
  - Cases = Confirmed Endometriosis
  - Controls = Confirmed Controls
  - Procedures = Procedure
- SCN.v1 - Surgically-Confirmed Endometriosis vs All excluding “ambiguous cases”
  - Cases = Confirmed Endometriosis
  - Controls = Non-Cases Except for “Ambiguous Cases”
  - Procedures = Surgery
- SCN.v2 - Surgically-Confirmed Endometriosis vs “confirmed controls”
  - Cases = Confirmed Endometriosis
  - Controls = Confirmed Controls

Procedures = Surgery
